# Safety and tolerability of RJx-01 to maintain muscle integrity in patients following hospitalisation with an acute exacerbation of COPD (MINT-COPD): protocol for a phase IIa randomised double-blind placebo-controlled trial

**DOI:** 10.64898/2026.11.06.26367435

**Authors:** Iram N. Qureshi, George Mills, Hamish J. C. McAuley, Lorna Latimer, Luc Meuleners, Ludo Haazen, Luke Baker, Phoebe Ashley, Niveditha Sathyanarayana, Emmanuel Oladipupo, Jill Clanchy, Ana C. Ugarte, Simon Verheijden, Ann Beliën, Jeroen Aerssens, Neil J. Greening

**Affiliations:** Institute for Lung Health, NIHR Leicester Biomedical Research Centre, Glenfield Hospital, Leicester, UK; Division of Respiratory Sciences, School of Medical Sciences, University of Leicester, Leicester, UK; Rejuvenate Biomed NV, Diepenbeek, Belgium

**Keywords:** COPD, Exacerbation, Muscle, Sarcopenia, Clinical Trial

## Abstract

**Introduction:** Chronic Obstructive Pulmonary Disease (COPD) is one of the leading causes of global disability and disability-adjusted life years. One of the key systemic features of COPD is impaired skeletal muscle mass and function, termed *sarcopenia*. The recognition of skeletal muscle dysfunction as a treatable trait in COPD opens a therapeutic avenue for pharmacological development, with the aim of improving physical function and reducing healthcare utilisation. A novel combination drug, RJx-01, was identified as a candidate therapy for sarcopenia based on preclinical evidence. The primary aim of this study is to evaluate the safety and tolerability of RJx-01 compared with placebo following hospitalisation with an acute exacerbation of COPD (AECOPD), when acute deterioration of skeletal muscle integrity is observed. Exploratory efficacy analyses will assess differences in muscle function, mass, and multi-omic-derived outcomes.

**Methods and analysis:** The MINT-COPD trial is a phase IIa, single centre, randomised controlled trial (RCT). Eighty participants admitted to hospital with AECOPD will be randomly assigned 1:1 to receive either oral RJx-01 or placebo, once daily for 24 weeks. The primary outcome is safety and tolerability of RJx-01 compared with placebo in COPD patients. Exploratory objectives aim to assess the preliminary effects of AECOPD-associated hospitalisation and RJx-01 treatment on muscle mass, function, and fatiguability, as well as patient reported symptoms and frailty. The optional skeletal muscle biopsy sub-study aims to assess the effects of AECOPD on proteomic and transcriptomic markers of skeletal muscle.

**Ethics and dissemination:** Ethical approval for this study was granted on 28-MAR-2025 by the East Midlands - Leicester Central Research Ethics Committee (ref: 25/EM/0032). The MINT-COPD trial commenced recruitment in May 2025 and aims establish the safety profile of RJx-01 in a population of individuals admitted to hospital with AECOPD. The results of the trial will be disseminated through publication in relevant peer-reviewed journals.

**What is already known on this topic:** Skeletal muscle dysfunction in COPD independently predicts mortality, readmission and disability, and hospitalisation for an acute exacerbation causes further muscle loss that does not immediately recover. Exercise-based rehabilitation may help but is completed by fewer than 10% of patients after an exacerbation.

**What this study adds:** RJx-01, a fixed-dose combination of metformin and galantamine, has improved muscle function in preclinical ageing models and attenuated disuse-related strength loss in healthy older adults. This Phase IIa study is the first to test its safety and tolerability in people hospitalised with an exacerbation of COPD.

**How this study might affect research, practice or policy:** The findings will establish whether treatment with RJx-01 is feasible in an acutely unwell, multimorbid population. An acceptable safety profile would provide the basis for an adequately powered efficacy trial and support further evaluation of skeletal muscle as a treatable trait in COPD.

**Trial registration:** ISRCTN14321370; Registered 09-APR-2025, https://www.isrctn.com/ISRCTN14321370

## Structured summary (1b)

**Table 1.**
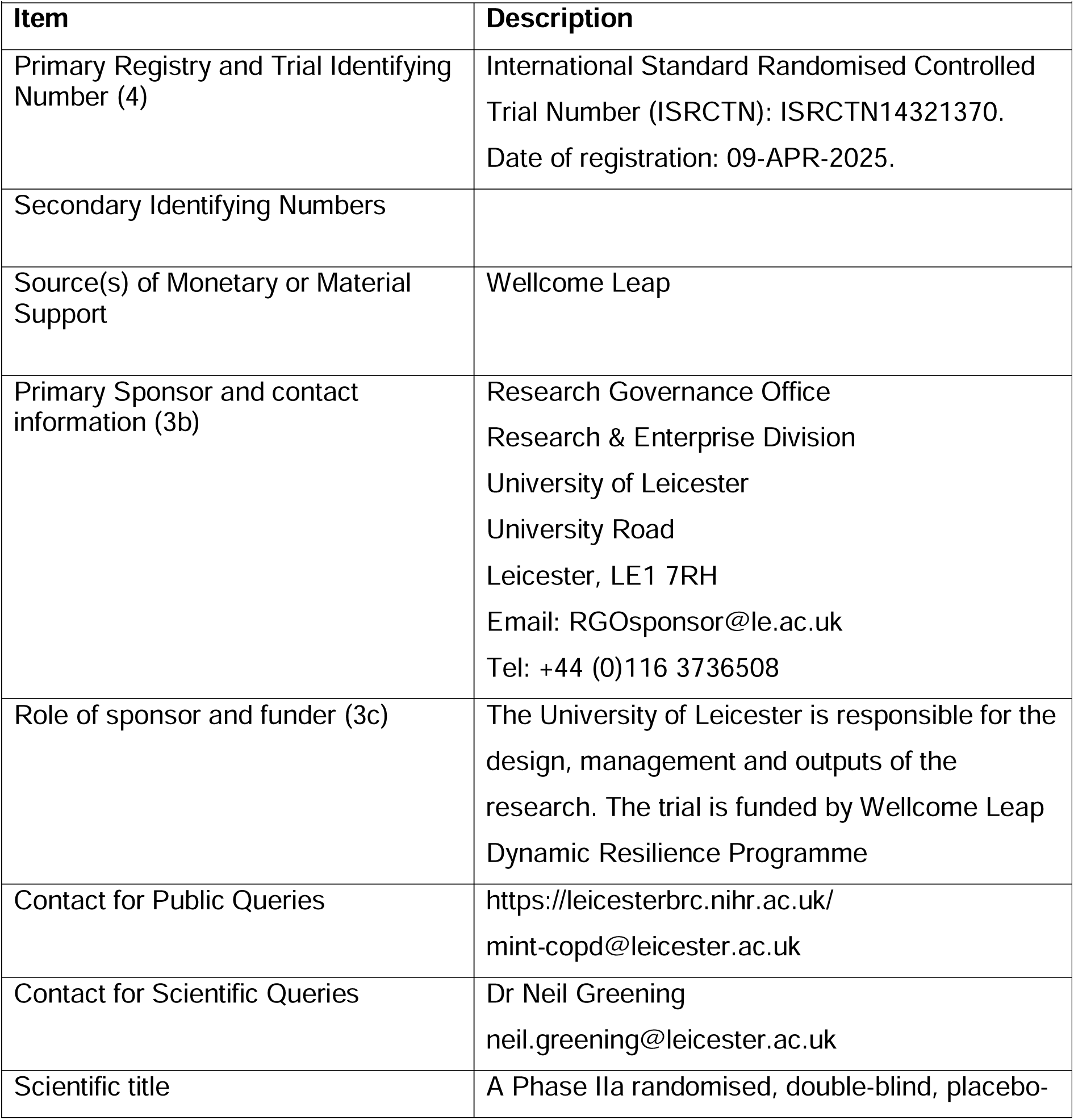
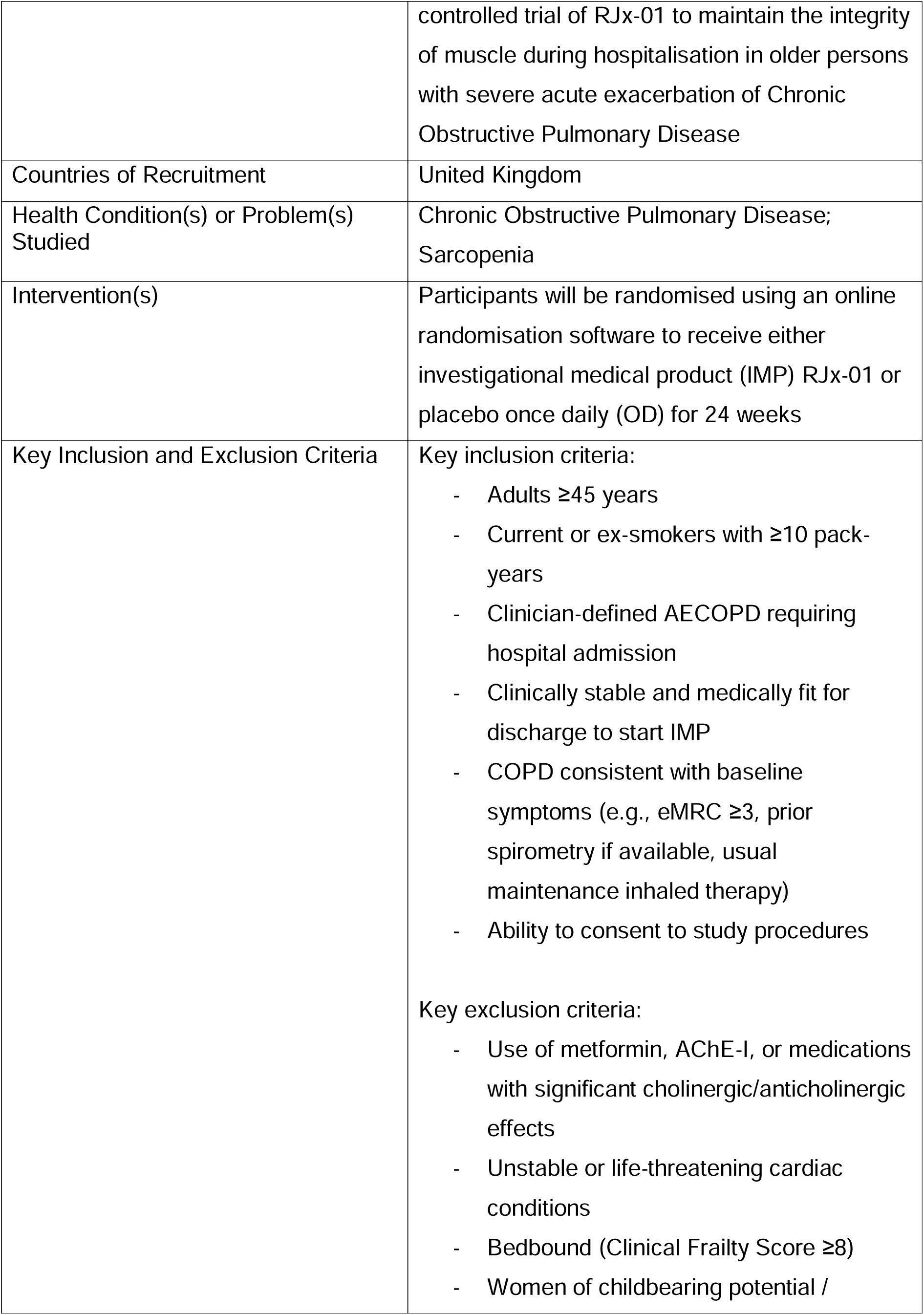
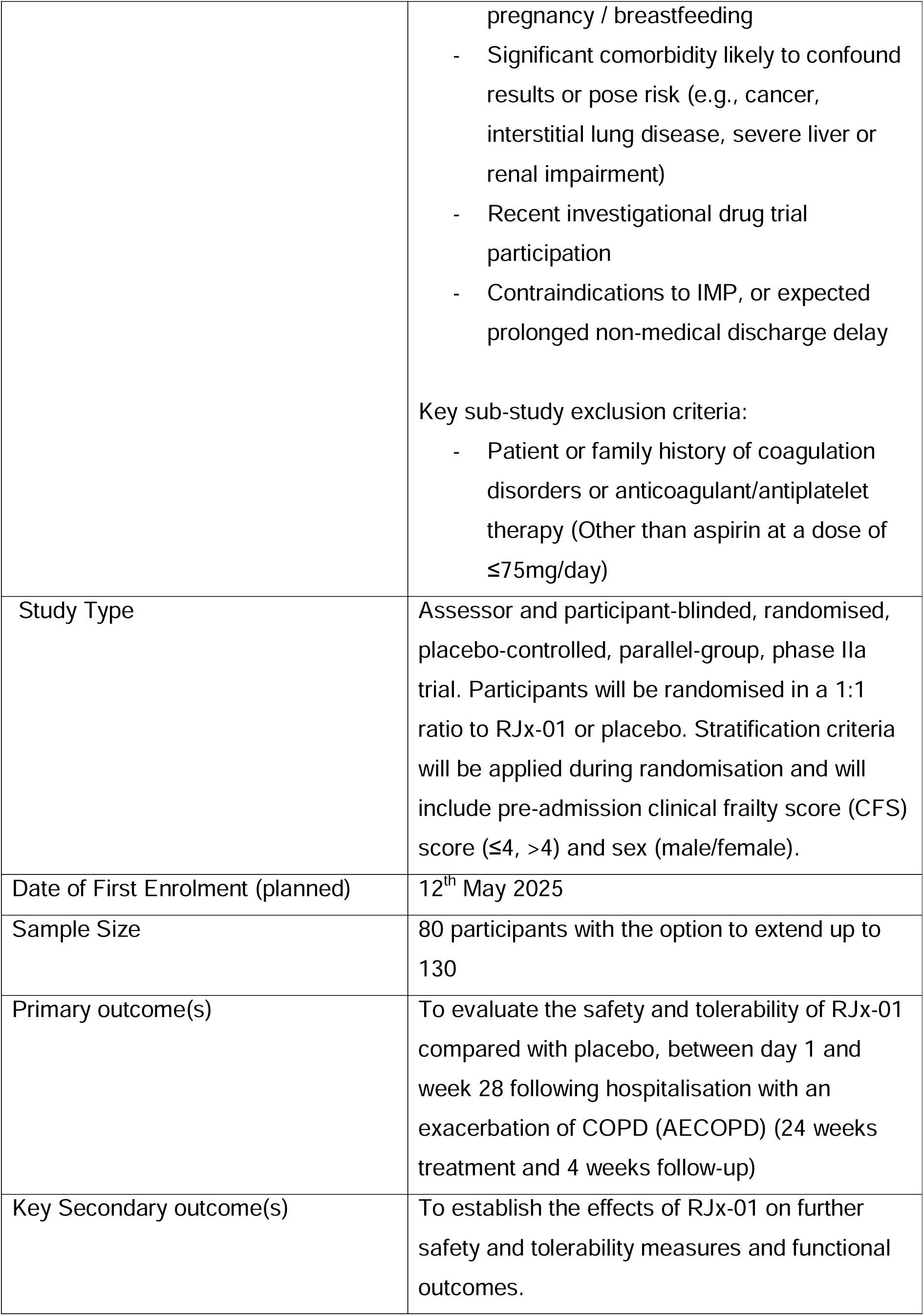
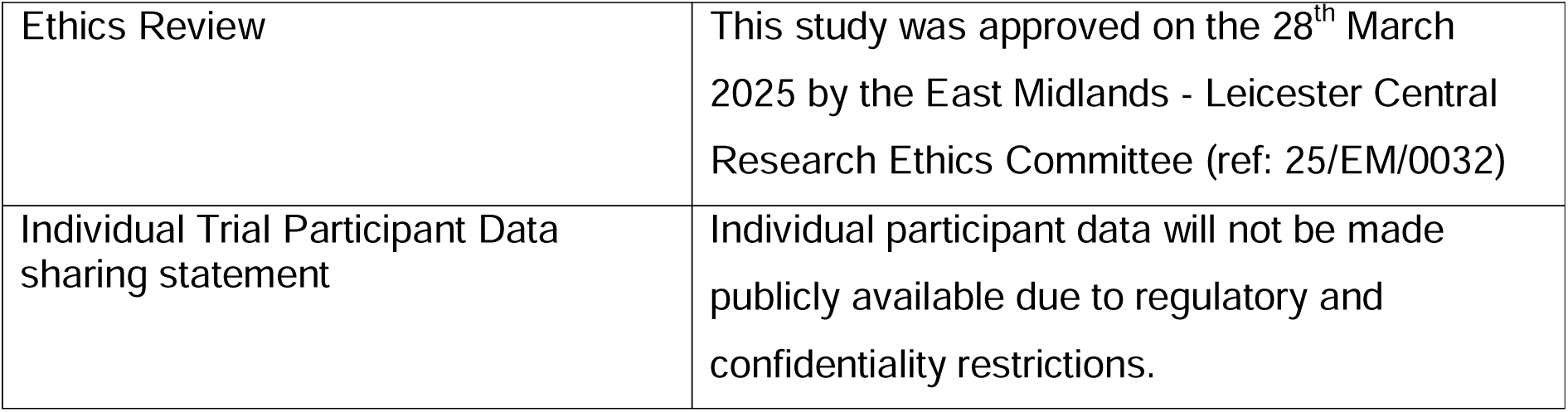
Structured summary of the MINT-COPD trial

## Introduction

### Background and rationale (9a)

Long-term conditions affect nearly one third of the UK population and cause 75% deaths globally, (1) with the number of people living with major illness projected to increase by over a third by 2040. (2) These conditions account for 70% of hospital bed days and healthcare spending. (3)

Many long-term conditions are linked to persistent disability, with skeletal muscle impairment prevalent due to inactivity and hospitalisation. (4, 5) In some individuals, these changes contribute to sarcopenia, a skeletal muscle disorder characterised by accelerated muscle strength and mass loss. (6) One of the primary co-existing conditions associated with sarcopenia is Chronic Obstructive Pulmonary Disease (COPD), a heterogeneous condition which ranks among the leading causes of mortality and disability globally. (7, 8) Increasing emphasis is being placed on identifying clinically relevant ‘treatable traits’ of COPD, (9) with skeletal muscle impairment representing a viable extrapulmonary target.

Individuals with COPD often experience flare ups termed ‘acute exacerbations’ (AECOPD). Severe exacerbations are associated with hospitalisation and death. Of those hospitalised, the 90-day mortality rate is 12%, (10) and nearly one in two are readmitted within 90-days in the UK. (11)

Skeletal muscle impairment is a strong predictor of mortality, morbidity, and healthcare utilisation in COPD, independent of the severity of lung impairment. (12–14) This impairment preferentially affects the muscles of locomotion and may be further exacerbated during hospitalisation by prolonged inactivity, tissue hypoxia, systemic inflammation and corticosteroid treatment. (13, 15, 16) Hospitalisation with AECOPD has been linked to reductions in quadriceps thickness that do not fully recover by 3 months post-discharge, (17) with reduced muscle mass predicting readmission risk. (12)

Post-exacerbation pulmonary rehabilitation improves physical function and reduces re- hospitalisation risk, however fewer than 10% of patients complete it. (18) The largest trial of exercise during the exacerbation itself found it could not be delivered with sufficient intensity to confer long term benefit. (19) Regardless, skeletal muscle in COPD retains plasticity in response to anabolic therapy such as exercise. (20–22) Skeletal muscle therefore appears to be a viable target in COPD for reducing downstream effects on mortality, disability and healthcare utilisation.

RJx-01, a novel fixed-dose combination of metformin hydrochloride (MET) and galantamine hydrobromide (GAL), is being developed to prevent and treat sarcopenia. Preclinical investigations indicated synergistic beneficial effects on muscle quality. (23) In *Caenorhabditis elegans*, RJx-01 enhanced lifespan, locomotion and muscle fibre integrity beyond either component alone. RJx-01 also improved physical performance and muscle strength in both *Opa1* knockout mice, which display accelerated sarcopenic features, and 22-month-old wild-type mice. (23)

In a Phase I trial in healthy older adults undergoing experimental lower extremity immobilisation, RJx-01 was safe and well tolerated. (24) Exploratory assessments indicated that RJx-01 attenuated declines in muscle strength and fatigue resistance and facilitated subsequent recovery. These early data provide mechanistic rationale for investigating RJx-01 in COPD-related sarcopenia.

### Explanation for the choice of comparator (9b)

To date, no pharmacological therapy is licensed to reduce physical or functional decline or reduce the risk of sarcopenia. Therefore, the intervention in this study will be provided solely as an adjunct to participants’ usual care, with no active comparator. A placebo group is required to address potential intervention biases both among the participants and the investigators.

### Objectives (10)

The primary objective is to evaluate the safety and tolerability of RJx-01 versus placebo between day 1 and week 28 following hospitalisation with AECOPD (24 weeks’ treatment and 4 weeks’ follow-up). Outcomes include Common Terminology Criteria for Adverse Events (CTCAE) grade ≥3 adverse events (AEs) and serious adverse events (SAEs) at least possibly related to the investigational medicinal product (IMP), all-cause hospital admissions, and mortality.

Exploratory objectives will assess effects on further safety and tolerability outcomes, physical function, muscle strength, fatiguability, muscle mass, quality of life, frailty status, biomarkers, lung function, physical activity, mortality, and healthcare utilisation.

A muscle biopsy sub-study will explore skeletal muscle multi-omics and structure in a subset of participants.

## Methods and analysis

### Methods: Patient and public involvement, and trial design Patient and public involvement (11)

Patients and the public contributed substantially to trial design. A focus group was held in August 2024 with the Centre for Exercise and Rehabilitation Science Patient and Public Involvement (PPI) group (Leicester, UK), including individuals with COPD and healthy volunteers. This informed the design and burden of outcome measures. One-to-one consultations were also held with individuals previously hospitalised for AECOPD.

Feedback led to a modified dual-energy x-ray absorptiometry (DXA) protocol for individuals who may struggle to lie flat, and a short magnetic resonance imaging (MRI) protocol to minimise discomfort. PPI also informed participant-facing materials, consent forms and transport arrangements. PPI will continue throughout the trial, informing dissemination.

### Trial design (12)

This is an assessor and participant-blinded, randomised, placebo-controlled, parallel- group, phase IIa clinical trial. Participants will be randomised using a 1:1 ratio to either RJx-01 or placebo. The trial design follows an exploratory, superiority framework focused on determining safety and tolerability, with exploratory evaluation of preliminary efficacy.

### Methods: Participants, interventions and outcomes Trial setting (13)

The single-centre trial will be conducted at Glenfield Hospital, University Hospitals of Leicester NHS Trust, Leicester, UK. Participants will be recruited following admission to the Clinical Decisions Unit (CDU) with a primary clinician-defined diagnosis of an acute exacerbation of COPD. Table 2 lists the characteristics of the people needed for this trial.

**Table 2.** Characteristics of the people who are needed for the trial

| Characteristic | The people we would expect to see included |
|---|---|
| Age | Adults aged ≥45 years are eligible for inclusion.<br>We expect the enrolled population to reflect the typical age distribution of hospitalised patients with AECOPD, which in national audit data shows a median age of 73 years (IQR 65–79). (10) |
| Sex | Individuals of any sex (male, female, or intersex) are eligible for inclusion. Sex will be recorded as reported by participants at enrolment. |
| Gender | Gender will be recorded as self-reported using the categories: male (including trans man); female (including trans woman); non-binary; other (not listed); not known; and not stated. |
| Race, ethnicity and ancestry | Race and ethnicity will be collected as a self-reported variable using the categories included in the study case report form. In the final trial report, race will be presented exactly as recorded, without aggregation of categories. |
| Socioeconomic status | Socioeconomic status (SES) is not collected in this study. No SES-specific indicators (e.g., deprivation indices, income bands, occupation, or education level) are captured in the CRF, and SES cannot be inferred from other recorded variables. |
| Geographic location | Participants will be recruited from Leicester, a predominantly urban area, with the study population reflecting a wider catchment that includes suburban and semi-rural communities across Leicestershire. As this is a single-site trial, specific geographical grouping of participants will not be reported. |
| Other characteristics relevant to the trial | In addition to demographic variables, disease-related characteristics including FEV1/FVC ratio, mMRC category, smoking history, and exacerbation history (e.g. prior hospitalisations for AECOPD). Comorbidities (excluding COPD) will be reported categorically. |

### Eligibility criteria for participants (14a)

Inclusion and exclusion criteria for the main trial and skeletal muscle sub-study are detailed in table 3.

**Table 3.** Eligibility criteria for participants

| **Inclusion criteria** |  |
|---|---|
| 1. | Symptoms typical of COPD when stable, defined as:<br>1.1. Baseline extended MRC dyspnoea grade ≥3 when stable<br>1.2. FEV1/FVC ratio <0.7 (if previous spirometry available)<br>1.3. Usually on maintenance inhaled therapy for COPD (LAMA, LABA ± ICS) |
| 2. | Clinician-defined acute exacerbation of COPD requiring hospital admission and change in treatment (e.g. systemic corticosteroids ± antibiotics). |
| 3. | Able to initiate the first dose of the investigational medicinal product (IMP) when medically fit for discharge. |
| 4. | Current or ex-smoker with ≥10 pack-years. |
| 5. | Age ≥45 years at screening. |
| 6. | Predicted hospital length of stay ≥48 hours. |
| 7. | Willing and able to provide informed consent. |
| 8. | Able to understand written/spoken English sufficiently to complete study measures, or supported by an English-speaking family member. |
| 9. | If male with a partner who is a woman of childbearing potential, willingness to comply with protocol contraception requirements. |
| **Exclusion criteria** |  |
| 1. | Use of, or contraindication to, metformin, any acetylcholinesterase inhibitor, or other systemic medications with significant cholinergic/anticholinergic effects within 6 months, except LAMA inhalers. |
| 2. | Known hypersensitivity to MET, GAL or any excipients, including tartrazine (FD&C yellow #5). |
| 3. | Hospitalisation within 2 weeks prior to first day of current hospital admission. |
| 4. | Three or more emergency/unplanned hospital admissions in the previous 6 months. |
| 5. | Unstable or life-threatening cardiac disease (e.g. myocardial infarction or unstable angina) within 12 months, excluding type 2 myocardial infarction. |
| 6. | Unstable or life-threatening arrhythmia requiring intervention in the last 3 months, QTc >500 ms, 2nd/3rd-degree bundle branch block, or high risk of QTc prolongation. |
| 7. | Congestive cardiac failure as the primary cause of dyspnoea/limitation. |
| 8. | Normally bedbound (Clinical Frailty Score ≥8). |
| 9. | Women of childbearing potential, or currently pregnant/breastfeeding. |
| 10. | Treatment considered palliative (life expectancy <3 months). |
| 11. | Anticipated delay in discharge due to social issues. |
| 12. | Other significant conditions likely to affect the trial (e.g. active cancer, interstitial lung disease, primary pulmonary hypertension, acute circulatory dysfunction, chronic liver disease, substance misuse). |
| 13. | Participation in another interventional trial within 3 months or use of an investigational medicinal product within 3 months or 5 half-lives. |
| 14. | ALT >3× upper limit of normal. |
| 15. | Severe renal impairment (CrCl <30 mL/min by Cockcroft–Gault). |
| 16. | Recent (<3 months prescription; <2 weeks non-prescription) use of drugs affecting muscle mass (e.g. testosterone, anti-androgens, anti-oestrogens, growth hormone, megestrol). |
| 17. | 20 mg/day prednisolone (or equivalent) for >28 days immediately before screening. |
| 18. | Recent (<3 months) treatment with an immunosuppressant (except corticosteroids). |
| **Sub-study inclusion criteria** |  |
| 1. | Meets all main trial eligibility criteria and has consented to MINT-COPD. |
| 2. | Willing and able to consent to the sub-study. |
| **Sub-study exclusion criteria** |  |
| 1. | Impaired clotting (history or family history of coagulopathy). |
| 2. | Current use of warfarin, therapeutic-dose LMWH, or any DOAC. |
| 3. | Current use of antiplatelet agents other than aspirin <75 mg/day. |

### Who will take informed consent? (32a)

Informed consent will be obtained by a trained study medic or appropriately qualified non- medical professional authorised on the delegation log. Potential participants will first be approached by their clinical care team. A study team member will then provide interested individuals with the Participant and Summary Information Sheets and answer questions. Written informed consent will be obtained and countersigned by the delegated study team member.

### Additional consent provisions for collection and use of participant data and biological specimens (32b)

Participants will consent to the collection, storage and analysis of samples (including DNA analysis) and use of de-identified data and images. De-identified data and samples may be shared with academic or commercial partners for trial-related analyses. Optional consent will be sought for future ethically approved research use of de-identified data, tissue banking of leftover samples, future genetic analysis, contact about future studies and a results summary.

## Intervention and comparator

### Intervention and comparator description (15a)

#### Investigational Medicinal Product (IMP)

Participants in the IMP (active) arm of the trial will receive RJx-01 (Rejuvenate Biomed NV, Diepenbeek, Belgium) via oral self-administration, once daily for 24 weeks. The trial IMP consists of a combination of metformin hydrochloride (MET) modified release, an antihyperglycemic agent that improves insulin sensitivity and reduces hepatic glucose production, and galantamine hydrobromide (GAL) immediate release, a reversible acetylcholinesterase inhibitor that increases central cholinergic neurotransmission.

The IMP will be supplied in pre-labelled kits by the hospital pharmacy, with written dosing and storage instructions. The first dose will be given when participants are medically fit for discharge, with subsequent doses self-administered at home. A low dose will be taken for 7 days to reduce gastrointestinal and cholinergic side effects, increasing to a maintenance dose after face-to-face medical review. Safety and tolerability will be monitored at monthly visits and by telephone.

#### Placebo (comparator)

The placebo comprises pre-labelled treatment kits containing inactive ingredients only, matched to the IMP in appearance, packaging, and labelling. Placebo kits will be provided to the clinical trials pharmacy team and dispensed using the same randomisation process, dosing schedule (including the initial titration period), administration instructions, and storage conditions as the IMP. All study staff involved in the dispensing of IMP and collection of outcome measures will remain blinded to treatment allocation.

### Criteria for discontinuing or modifying allocated intervention/comparator (15b)

The criteria for permanent or temporary discontinuation of the intervention are listed in table 4.

**Table 4.** Criteria for discontinuing the intervention

| **Criteria for permanently discontinuing the intervention include:** |  |
|---|---|
| 1) | Reduced creatinine clearance <30 mL/min and lactic acidosis defined as an anion gap >12 mEq/L and deemed at least possibly related to allocated treatment. |
| 2) | Serious adverse event deemed at least possibly related to allocated treatment. |
| 3) | QTc >500 ms OR uncorrected QT >600 ms |
| 4) | Increase from baseline QTc of >60 ms |
| 5) | QTc ≥530 ms if baseline QTc 450–480 ms with bundle branch block. |
| 6) | Threat to participant’s safety or well-being in the view of the investigator |
| **Intervention may be temporarily discontinued at the discretion of the investigator. For example, in the case of:** |  |
| 1) | General anaesthesia |
| 2) | Respiratory acidosis |
| 3) | Severe dehydration |
| 4) | Use of iodinated contrast agents |
| 5) | Renal failure |
| 6) | Swallowing difficulties |

### Strategies to improve adherence to intervention/comparator (15c)

When recruited to the trial, participants will be given a diary to complete at home and asked to record if they took the medication on each day and at what time. The participants will then be asked to bring this diary to subsequent visits. When participants attend for study visits, they will be asked to bring in with them all empty and full sachets so the site staff can perform accountability checks. Additionally, if the participants are admitted to the hospital during the time of the trial, the IMP may be re-prescribed and additional boxes provided.

### Concomitant care permitted or prohibited during the trial (15d)

Participants may not be involved in other interventional clinical trials whilst participating in this study. All other necessary care is allowed. Concomitant medications, treatments will be monitored throughout the study – with pulmonary rehabilitation uptake reported as an exploratory outcome measure between groups.

#### Ancillary and post-trial care (34)

There is no provision for any ancillary or post-trial care as part of the trial. The host centre does not provide compensation; participants would be able to follow NHS channels for complaints.

### Outcomes (16)

The joint primary outcomes of this trial consist of: the incidence of at least possibly related AEs of CTCAE grade ≥3 (CTCAE v5.0 - November 27, 2017), at least possibly related SAEs, hospital admissions (for any reason), and mortality rate. Relatedness is according to the investigator’s opinion. Total incidence will be compared between IMP and placebo groups (24 weeks treatment and 4 weeks follow-up) between day 1 and week 28 following hospitalisation with AECOPD to determine safety. Exploratory outcomes encompassing the domains described in section 10 are listed in the supplementary material, with examples of variables that may be reported for each outcome category. Specific variables and analysis methods will be pre-specified in the Statistical Analysis Plan

### Skeletal muscle sub-study

A skeletal muscle biopsy sub-study will be conducted in a subset of MINT-COPD participants to elucidate the cellular mechanisms underpinning changes in skeletal muscle function and mass during and after hospitalisation for an acute exacerbation of COPD, and – if sample size permits – to assess the impact of RJx-01 during this period. Percutaneous vastus lateralis biopsies will be obtained at baseline (during the index hospital admission), and at approximately 8 and 16 weeks post-discharge. Tissue will undergo comprehensive cellular and molecular analysis including transcriptional profiling, proteomics, immunohistochemistry, and electron microscopy to characterise the skeletal muscle phenotype. Participants enrolled in the sub-study will be randomised as part of the main trial.

### Harms (17)

Preclinical and phase 1 data show no new or increased safety findings attributable to combined MET and GAL. RJx-01 was well tolerated in healthy older adults, with no product-related serious adverse events (SAEs) during the DSUR period.

Pre-planned hospital admissions and COPD related events such as exacerbations are expected in this population and will be reported but not considered AEs/SAEs.

Expected procedural harms include mild discomfort from blood draws and muscle soreness from strength testing. Muscle biopsies (sub-study only) may cause local bleeding, bruising, pain, or haematoma; infection or significant bleeding is uncommon. AEs will be assessed at each visit, and participants will be encouraged to report events by telephone between visits. SAEs will be reported within 24 hours. AEs will be coded using MedDRA and grouped by System Organ Class.

### Participant timeline (18)

Figure 1 depicts participant flow during the trial.

**Figure 1.**
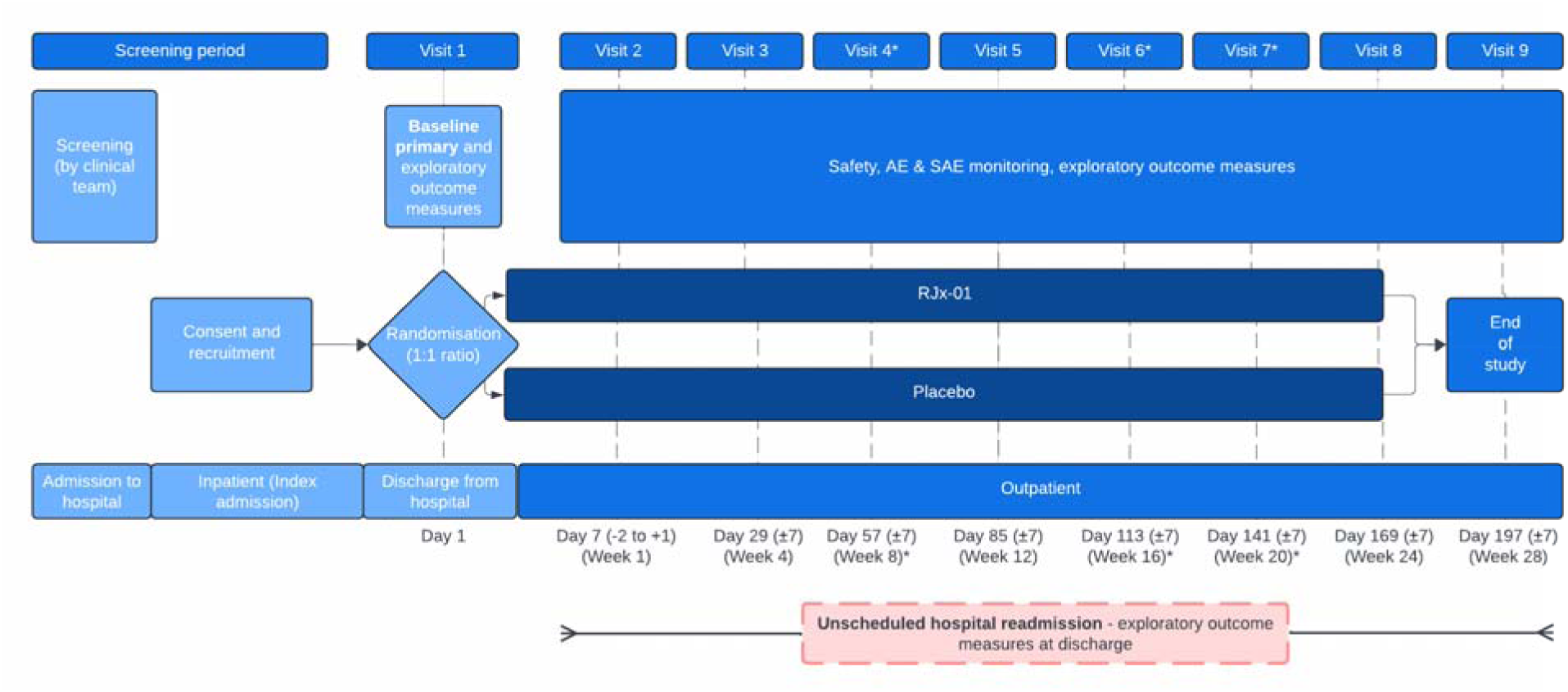
Participant study timeline. *Optionally as telephone visits (visit 4, 6, and 7)

### Sample size (19)

With limited data on RJx-01 and none in this population, a formal power calculation was not possible. At least 40 participants per arm may provide initial insights into the primary safety and exploratory efficacy endpoints. The study therefore aims to randomise at least 80 participants (40 per treatment arm), with the option of up to 130 participants (65 per treatment arm) as a larger sample would benefit the trial.

### Recruitment (20)

Glenfield Hospital (University Hospitals of Leicester NHS Trust) has over 2,000 AECOPD admissions annually. Previous inpatient AECOPD trials, including drug trials, have recruited approximately 10–20 participants monthly. Potential participants will be identified via the hospital’s Clinical Decisions Unit as soon as practicable after admission and approached by their healthcare team.

## Assignment of interventions: randomisation

### Sequence generation: who will generate the sequence (21a)

The random allocation will be generated by a third-party web-based randomisation system. Members of the trial team involved in study assessments and participants will be blinded to treatment assignment.

### Sequence generation: type of randomisation (21b)

Participants will be randomised in a 1:1 ratio to RJx-01 or placebo using restricted randomisation with stratification by pre-admission Clinical Frailty Scale score (≤4 versus >4) and sex at birth (male/female).

### Allocation concealment mechanism (22)

Allocation will be concealed using a centralised, third-party web-based randomisation system. The allocation sequence and unblinded data will be accessible only to the independent Data Safety Monitoring Committee (DSMC) statistician (section 28a) and designated unblinded personnel.

### Implementation (23)

Trained, blinded personnel will enrol participants and enter eligibility, pseudonymised identification and stratification data into the system, which will assign treatment and a unique randomisation ID. Kit numbers will be emailed to relevant personnel, including pharmacy.

### Assignment of interventions: blinding Who will be blinded (24a)

Participants, investigators, pharmacy, and all involved in trial conduct or with any other interest in this trial will remain blind to the randomised treatment assignments until after database lock.

### How will blinding be achieved (24b)

Both RJx-01 and placebo are packaged identically and are visually indistinguishable. Kit numbers will be used to ensure all individual treatment doses can be attributed to their respective allocation instance on the electronic database. The treatment group (IMP / placebo) associated with each kit is held securely within the web-based randomisation system with access unavailable to all members of the study team, except in the case of emergency unblinding.

### Procedure for unblinding if needed (24c)

Unblinding will only occur in the case of a medical emergency where knowledge of the allocated treatment is needed for clinical care. As there is no specific antidote for RJx-01 or its components, management would be primarily supportive, so unblinding would rarely alter immediate treatment. Emergency unblinding will be available via the audit-traceable web-based randomisation system, authorised by the PI or a delegated Co-Investigator.

## Data collection and management

### Plans for assessment and collection of outcomes (25a)

All outcome assessments (summarised in the supplementary material) will follow standard operating procedures. Specialist assessments (e.g. isokinetic dynamometry, DXA, MRI) will be performed by qualified operators using calibrated equipment. To minimise inter- rater variability, assessors will complete study-specific training and, where feasible, one individual will pre-process all data for each measure (e.g. DXA region of interest).

Validated patient-reported outcome measures will be completed before other assessments at each visit. Physiological and functional assessments will follow standardised protocols to ensure consistency.

Laboratory samples will be handled according to a site laboratory manual. Clinical data will be recorded on electronic case report forms with built-in range checks, logic rules and mandatory fields.

### Plans to promote participant retention and complete follow-up (25b)

Participants will be encouraged to attend all visits face-to-face to ensure completeness of outcome measures. However, visits 4, 6, and 7 will be offered as optional telephone appointments for participants already established on treatment and for whom no outstanding safety concerns exist warranting face-to-face follow-up. To encourage continued participation in those who withdraw from study treatment, all further study assessments (excluding a 28-day post-discontinuation safety follow-up visit) will be optionally conducted via telephone.

### Data management (26)

Participants will be assigned a unique study identification number at consent, used on all study documents in place of identifiable information. Trained staff will enter data into an electronic case report form (eCRF) developed by a third party, with in-system query management. Source data will be verified during monitoring visits in accordance with ICH- GCP. A data management plan will be developed before first enrolment.

### Confidentiality (33)

Data will be handled in accordance with the UK Data Protection Act, EU General Data Protection Regulation, ICH-GCP and UK Policy Framework for Health and Social Care Research. Data will be pseudonymised, with the key linking identification numbers to personal details held securely at site and accessible only to authorised staff. Identifiable data will be collected only where necessary, with explicit consent for its storage and sharing. Sponsor and regulatory representatives may access source data for monitoring, audit or inspection. The sponsor is the data controller.

## Statistical methods

### Statistical methods for primary and exploratory outcomes (27a)

The primary safety analysis will compare rates of IMP-related AEs of grade ≥3, IMP-related SAEs, IMP-related hospitalisations and deaths between treatment arms. Only events with onset on or after the first administration of the investigational medicinal product will be included; events with an unknown start date will be classified as treatment emergent. Treatment groups will be compared using a Cochran-Mantel-Haenszel test for general association, controlling for stratification factors.

For exploratory efficacy outcomes, continuous endpoints will be analysed using a mixed model for repeated measures with baseline value, stratification factors, treatment, time, and treatment-by-time interaction as fixed effects and participant as a random effect. Non- parametric alternatives will be used where distributional assumptions are not met. Within- group differences versus baseline will be tested using a paired t-test or Wilcoxon signed ranks test. Categorical endpoints will be compared using the Cochran-Mantel-Haenszel test for general association. Time-to-readmission will be summarised using Kaplan-Meier methods and compared using the stratified log-rank test.

### Who will be included in each analysis (27b)

The primary safety analysis will include all randomised participants who receive at least one dose of the IMP or placebo. Exploratory efficacy analyses will be conducted on the intent-to-treat population containing all randomised participants who receive at least one dose of the IMP or placebo and who have post-baseline data. A summary of participant flow including numbers screened, screen-failed, randomised, exposed, discontinued (with reasons), and completed will be reported in accordance with the CONSORT statement.

### How missing data will be handled in the analysis (27c)

The mixed model for repeated measures used for continuous exploratory endpoints assumes data are missing at random and uses all available observations without imputation. Pre-specified intercurrent event strategies will be applied: treatment policy for rehospitalisation, retaining data during and after readmission, and while-on-treatment for death and IMP discontinuation, using data up to the event. Sensitivity analyses under alternative missing data assumptions will be detailed in the statistical analysis plan.

### Methods for additional analyses (e.g. subgroup analyses) (27d)

Baseline demographics and clinical characteristics will be summarised by treatment arm using means, standard deviations, medians, and quartiles for continuous variables and frequencies and percentages for categorical variables. Concurrent treatments (pharmacological and non-pharmacological) and important protocol deviations will be tabulated by treatment arm. Any pre-specified subgroup or sensitivity analyses will be detailed in the statistical analysis plan, which will be finalised prior to database lock.

### Interim analyses (28b)

No formal interim analysis is planned for this trial outside of the safety analyses provided to the DSMC, which is prepared by an independent unblinded statistician.

### Protocol and statistical analysis plan (5)

The full study protocol is available upon request. A detailed standalone statistical analysis plan will be developed by the trial statistician and finalised prior to database lock.

## Oversight and monitoring

### Composition of the coordinating centre and trial steering committee (3d)

The trial will be coordinated day to day by the research team at the NIHR Leicester BRC. The Trial Management Group (TMG), comprising the Chief Investigator, Research Fellow(s), Trial Manager, and Trial statistician, will oversee operational aspects including protocol compliance, safety reporting, recruitment, and budget. The TMG will meet monthly and refer significant issues to the Trial Steering Committee (TSC) or Investigators.

The TSC will provide independent oversight, considering blinded data reports, and recommendations from the DSMC, to ensure compliance with the protocol, regulations, and Good Clinical Practice (GCP). The TSC will comprise an Independent Chairperson and two other independent members. The TSC will meet every six months; the Chief Investigator and Principal Investigator will attend without voting rights.

### Composition of the data monitoring committee, its role and reporting structure (28a)

The DSMC will consist of independent experts. The role of the DSMC is to assess participant safety and oversee the conduct of the clinical trial. Members of the DSMC will have no conflicts of interest in relation to the trial.

The DSMC will review blinded and/or unblinded safety data as appropriate. It will report its findings and make recommendations to the TSC. To ensure timely input into TSC oversight, DSMC meetings will be scheduled prior to each TSC meeting. The DSMC will meet every six months throughout the active phases of the trial, or more frequently at the discretion of the DSMC Chair, and will operate according to a DAMOCLES charter.

### Frequency and plans for auditing trial conduct (29)

The University of Leicester, as Sponsor, operates a risk-based monitoring and audit programme. A risk assessment will be conducted and a monitoring plan developed by the Sponsor in accordance with the level of risk identified to participant safety, trial integrity, and data validity. Regular monitoring will be conducted in accordance with ICH GCP guidelines.

### Protocol amendments (31)

Amendments will require Sponsor, ethics committee, competent authority and site approval before implementation. This manuscript reflects all amendments to date; subsequent amendments will be available via the trial registry.

### Dissemination policy (8)

A publication plan will be developed by the TMG, subject to approval by the Chief Investigator, sponsor and Rejuvenate Biomed NV. Results will be published in peer- reviewed journals, acknowledging all supporting organisations, with authorship following International Committee of Medical Journal Editors guidelines. Participants will be invited, where possible, to a dissemination event to learn the results.

## Discussion

Physical frailty and sarcopenia represent major clinical burdens in chronic respiratory disease, with consequences that extend beyond reduced quality of life and functional independence to poorer prognosis and elevated risk of re-hospitalisation. (12, 25) Previous pharmacological approaches targeting muscle size in COPD via anabolic pathways have failed to demonstrate concomitant improvements in functional capacity and health status. (26–28) Exploring alternative therapeutic pathways is therefore a clear next step.

RJx-01 provides a candidate novel approach to address a specific mechanistic pathway of muscle dysfunction occurring in COPD patients. Building on synergistic effects for skeletal muscle in preclinical models and in healthy older adults undergoing immobilisation, the combination – further supported by an optimised low dose formulation - is hypothesised to demonstrate a favourable safety and tolerability profile alongside a preliminary efficacy signal for skeletal muscle outcomes in patients hospitalised with AECOPD.

MINT-COPD will employ a two-arm, randomised, placebo-controlled design, as no pharmacological standard of care exists for skeletal muscle dysfunction in this setting. Whilst the primary objective is to evaluate safety and tolerability, robust physiological assessments will also explore preliminary effects on skeletal muscle integrity. Despite the absence of a formal sample size calculation, a recruitment target of 80 is deemed sufficient to detect signs of meaningful differences in many of the key strength and functional outcomes, based on effect sizes from previous interventional studies. (29–31) This trial will also represent one of the most detailed longitudinal assessments of skeletal muscle recovery following hospitalisation for AECOPD, providing unique insights into post- exacerbation recovery trajectories.

Whilst pulmonary rehabilitation is highly effective at reducing muscle weakness and frailty, there exists a participation paradox where frailty is itself a predictor of programme non- completion. (32) As current guidance encourages post-exacerbation pulmonary rehabilitation, rather than excluding participants undertaking it, we will monitor and report uptake and completion rates.

Overall, this population is particularly at risk of frequent hospital readmissions and incomplete physical recovery, with higher rates of sarcopenia compared to the general ageing population. The post-AECOPD period therefore provides a unique opportunity to intervene and address the cycle of muscle weakness, functional deterioration and escalating dependence on secondary care. Exploring the safety and tolerability of RJx-01 in this at-risk population is an important first step.

## Trial status

Recruitment began in May 2025 and is ongoing. With a 24-week treatment and 4-week follow-up period, last participant last visit is expected to occur approximately 7 months after the first visit of the last participant recruited.

## Abbreviations

AE: Adverse Event
AECOPD: Acute Exacerbation Of COPD
CDU: Clinical Decisions Unit
COPD: Chronic Obstructive Pulmonary Disease
CrCl: Creatinine Clearance
CRF: Case Report Form
CTCAE: Common Terminology Criteria for Adverse Events
CTIMP: Clinical Trial of an Investigational Medicinal Product
DSMC: Data and Safety Monitoring Committee
DXA: Dual-energy X-ray Absorptiometry
FEV_1_/FEV: Forced Expiratory Volume in 1 second / total Forced Expiratory Volume ratio
GAL: Galantamine Hydrobromide
GCP: Good Clinical Practice
ICMJE: International Committee of Medical Journal Editors
ICS: Inhaled Corticosteroids
IQR: Interquartile Range
IMP: Investigational Medicinal Product
LABA: Long-Acting Beta-2 Agonist
LAMA: Long-Acting Muscarinic Antagonists
LWMH: Low-Molecular-Weight Heparin
MET: Metformin Hydrochloride
mMRC: modified Medical Research Council dyspnea scale
MRI: Magnetic Resonance Imaging
OD: Once Daily
PPI: Patient and Public Involvement
QTc: Corrected QT Interval
SAE: Serious Adverse Event
SEM: Standardized Error of the Mean
SES: Socioeconomic Status
TEAEs: Treatment Emergent Adverse Events
TESAEs: Treatment Emergent Serious Adverse Events
TMG: Trial Management Group

## Declarations Acknowledgements

This work was supported by the NIHR Leicester Biomedical Research Centre and the NIHR Leicester Clinical Research Facility. We would like to thank the following individuals for their contributions to the protocol: Dr Sam Corveleyn, CMC Leader, Rejuvenate Biomed NV Dr Dieudonné Bwirire, Desirée Kanters, Griet Boogaerts, Clinical Operations Leaders, Rejuvenate Biomed NV Dr Matthew Richardson, Statistician, University of Leicester Professor Ken Smith, Professor of Metabolic Mass Spectrometry, University of Nottingham Professor Philip Atherton, Professor of Clinical, Metabolic & Molecular Physiology, University of Nottingham Dr Daniel Wilkinson, Principal Research Fellow, University of Nottingham

## Authors’ contributions (3a)

*INQ and GM are joint co-authors. NJG is the Chief Investigator; he conceived the study, led the proposal and protocol development. HJCM, JA, and AB contributed to study design and to the development of the proposal and protocol. INQ, GM, LL, LH, SV, and ACU contributed to study design and to the development of the protocol. LM contributed to study design, statistical planning, and to the development of the protocol. LB contributed to the design of the muscle biopsy sub-study and to the development of the protocol. GB, SV, PA, NS, and EO contributed to the review of the protocol. All authors read and approved the final manuscript.

Authorship on all future trial publications will be determined in accordance with the International Committee of Medical Journal Editors (ICMJE) criteria for authorship. Individuals who do not meet authorship criteria will be acknowledged appropriately. No artificial intelligence technologies were used in the preparation of this manuscript.

## Availability of data and materials (6)

The datasets generated and/or analysed during this trial will not be publicly available due to the clinically sensitive nature of the data collected. Access to the final trial dataset will be restricted to the Chief Investigator and named trial team members. Any requests for data access will be considered on a case-by-case basis and will be subject to applicable data protection legislation, institutional governance requirements, and contractual agreements between the sponsoring institution and trial partners.

## Ethics approval and consent to participate (30)

This study received ethical approval on 28-MAR-2025 by the East Midlands - Leicester Central Research Ethics Committee (2 Redman Place, Stratford, London, E20 1JQ, United Kingdom, ref: 25/EM/0032). Written, informed consent to participate will be obtained from all participants. A copy of informed consent forms for the main study and sub study are available on request.

## Consent for publication

Not applicable

## Competing interests (7b)

NJG reports personal and institutional grants and fees from Wellcome Leap, GlaxoSmithKline, Roche, AstraZeneca, Chiesi, Pulmonx, Roche, Sanofi and is chair of the British Thoracic Society COPD Specialist Advisory Group. LM, LH, ACU, SV, AB and JA were employed or funded by Rejuvenate Biomed at the time the study protocol was developed. AB holds patents related to RJx-01.

## Authors’ information (optional)

Not applicable

## Data Availability

Not applicable

