## supplementary material for "Safety and tolerability of RJx-01 to maintain muscle integrity in patients following hospitalisation with an acute exacerbation of COPD (MINT-COPD): protocol for a phase IIa randomised double-blind placebo-controlled trial"

### Exploratory Outcome Measures: MINT-COPD

*The following table provides an overview of exploratory outcome categories and illustrative examples of variables that may be reported. This is not an exhaustive list; specific variables will be pre-specified in the Statistical Analysis Plan.*

| Outcome Category | Examples of Variables | Timeframe |
| --- | --- | --- |
| Safety | Adverse events (TEAEs, TESAEs), vital signs (heart rate, blood pressure, temperature), ECG (QTc interval), blood biomarkers (liver function, haematology, metabolic panel). | Baseline and all study visits, including hospital readmissions |
| Physical Function | Short Physical Performance Battery (SPPB), quadriceps strength, handgrip strength, muscle fatigability measures. | Baseline and subsequent study visits |
| Body Composition | Body weight, lean mass and fat mass (bioelectrical impedance analysis), skeletal muscle mass (DXA), quadriceps muscle characteristics (MRI). | Baseline and subsequent study visits |
| Patient-Reported Outcomes | Respiratory specific quality of life (SGRQ-C), sarcopenia-related quality of life (SarQoL), sarcopenia risk (SARC-F). | Baseline and subsequent study visits |
| Frailty and Cognition | Clinical Frailty Scale (CFS), Fried's Frailty Phenotype, cognitive function (Mini-ACE). | Baseline and subsequent study visits |
| Physical Activity | Daily step count, time in moderate-to-vigorous physical activity, inactivity time (wearable accelerometry). | 7-day periods at baseline and subsequent study visits |
| Lung Function | FEV1, FVC, FEV1/FVC ratio (spirometry). | Selected study visits |
| IMP-Related Assessments | IMP acceptability (participant questionnaire), pharmacokinetic profile (blood sampling). | Selected study visits |
| Healthcare Utilisation | Time to hospital readmission, total hospital days, outpatient contacts, uptake to pulmonary rehabilitation, mortality. | Continuously throughout study period |
| Exploratory Biosamples | Blood (plasma, serum), urine, sputum, and muscle biopsy (vastus lateralis) samples for exploratory biomarker, transcriptomic, proteomic, and metabolomic analyses. | Baseline and selected study visits |
